# Digestive Manifestations in Patients Hospitalized with COVID-19

**DOI:** 10.1101/2020.07.07.20143024

**Authors:** B. Joseph Elmunzer, Rebecca L. Spitzer, Lydia D. Foster, Ambreen A. Merchant, Eric F. Howard, Vaishali A. Patel, Mary K. West, Emad Qayed, Rosemary Nustas, Ali Zakaria, Marc S. Piper, Jason R. Taylor, Lujain Jaza, Nauzer Forbes, Millie Chau, Luis F. Lara, Georgios I. Papachristou, Michael L. Volk, Liam G. Hilson, Selena Zhou, Vladimir M. Kushnir, Alexandria M. Lenyo, Caroline G. McLeod, Sunil Amin, Gabriela N. Kuftinec, Dhiraj Yadav, Charlie Fox, Jennifer M. Kolb, Swati Pawa, Rishi Pawa, Andrew Canakis, Christopher Huang, Laith H. Jamil, Andrew M. Aneese, Benita K. Glamour, Zachary L Smith, Katherine A. Hanley, Jordan Wood, Harsh K. Patel, Janak N. Shah, Emil Agarunov, Amrita Sethi, Evan L. Fogel, Gail McNulty, Abdul Haseeb, Judy A. Trieu, Rebekah E. Dixon, Jeong Yun Yang, Robin B. Mendelsohn, Delia Calo, Olga C. Aroniadis, Joseph F. LaComb, James M. Scheiman, Bryan G. Sauer, Duyen T. Dang, Cyrus R. Piraka, Eric D. Shah, Heiko Pohl, William M. Tierney, Stephanie Mitchell, Ashwinee Condon, Adrienne Lenhart, Kulwinder S. Dua, Vikram S. Kanagala, Ayesha Kamal, Vikesh K. Singh, Maria Ines Pinto-Sanchez, Joy M. Hutchinson, Richard S. Kwon, Sheryl J. Korsnes, Harminder Singh, Zahra Solati, Amar R. Deshpande, Don C. Rockey, Teldon B. Alford, Valerie Durkalski, the North American Alliance for the Study of Digestive Manifestations of COVID-19.

## Abstract

**Background:** The prevalence and significance of digestive manifestations in COVID-19 remain uncertain.

**Methods:** Consecutive patients hospitalized with COVID-19 were identified across a geographically diverse alliance of medical centers in North America. Data pertaining to baseline characteristics, symptomatology, laboratory assessment, imaging, and endoscopic findings from the time of symptom onset until discharge or death were manually abstracted from electronic health records to characterize the prevalence, spectrum, and severity of digestive manifestations. Regression analyses were performed to evaluate the association between digestive manifestations and severe outcomes related to COVID-19.

**Results:** A total of 1992 patients across 36 centers met eligibility criteria and were included. Overall, 53% of patients experienced at least one gastrointestinal symptom at any time during their illness, most commonly diarrhea (34%), nausea (27%), vomiting (16%), and abdominal pain (11%). In 74% of cases, gastrointestinal symptoms were judged to be mild. In total, 35% of patients developed an abnormal alanine aminotransferase or total bilirubin level; these were elevated to less than 5 times the upper limit of normal in 77% of cases. After adjusting for potential confounders, the presence of gastrointestinal symptoms at any time (odds ratio 0.93, 95% confidence interval 0.76-1.15) or liver test abnormalities on admission (odds ratio 1.31, 95% confidence interval 0.80-2.12) were not independently associated with mechanical ventilation or death.

**Conclusions:** Among patients hospitalized with COVID-19, gastrointestinal symptoms and liver test abnormalities were common but the majority were mild and their presence was not associated with a more severe clinical course

## Introduction

Even though COVID-19 is primarily a respiratory illness, the digestive system has been implicated in disease expression, transmission, and possible pathogenesis. The responsible virus – Severe Acute Respiratory Syndrome Coronavirus 2 (SARS-CoV-2) – gains cellular entry through the Angiotensin Converting Enzyme 2 (ACE2) receptor, which is present in the gastrointestinal tract at higher levels than in the respiratory system (1-3). Viral RNA has been detected in the stool of approximately 50% of affected patients (1,4-5), and it is hypothesized that enteric infection might modulate the severity of pulmonary and systemic illness through alterations in the microbiome, dysregulated intestinal immunity, and/or increased gut permeability (6-8).

Digestive manifestations may be common in patients with COVID-19, although reports are conflicting, and the true prevalence remains uncertain. Early series from China and two recent meta-analyses suggest that gastrointestinal symptoms occur in less than 10% of patients (9-13) whereas other studies demonstrate proportions in the range of 30-60% (14-17). The prevalence of abnormal liver tests similarly varies from 15 to 50% (9,12,13,15). More importantly, the significance of digestive manifestations in COVID-19, in terms of impact on the alimentary tract and liver, and on overall outcomes, is unknown. The presence and magnitude of gastrointestinal symptoms and hepatic abnormalities have mirrored disease severity in some studies, but this observation has been inconsistent (18-21).

Reports on the digestive manifestations of COVID-19 have been limited in scope, reflecting the experience of a single hospital or isolated geographic region, and have used varying and potentially biased sampling strategies. We aimed to systematically and rigorously assess the prevalence, spectrum, and severity of digestive manifestations in consecutive patients hospitalized with COVID-19 across geographically diverse medical centers in North America. We also explored the association between the presence of digestive manifestations and overall outcomes.

## Methods

### Study design

This was an observational cohort study conducted through an alliance of 36 medical centers in the United States and Canada. Institutional review board approval was obtained at each center prior to patient identification and data collection.

### Patients

Adult patients who were hospitalized with a confirmed diagnosis of COVID-19 according to local testing standards were considered eligible. To ensure an unbiased sample, we aimed to enroll the first 50-100 consecutive patients meeting eligibility criteria at each participating institution. Potentially eligible patients were identified by site investigators using multiple methods, including but not limited to, data warehouse queries, electronic research subject identification tools, and lists provided by the infectious diseases service or other relevant hospital entities.

### Data collection and quality assurance

Demographic, clinical, laboratory, radiographic, and endoscopic data from symptom onset until discharge or death were manually abstracted through review of electronic health records by study personnel under the oversight of a designated clinician-investigator. Deidentified data were entered directly into an electronic data collection form (**Supplementary Appendix**). When patients had not been dispositioned by the end of the study period, data were collected within 3 days of study closure.

Data quality was ensured to the greatest extent possible using a three-tiered system. First, formal instructions and consistent communications between the data coordinating center and co-investigators emphasized the importance of ensuring response accuracy at the site level and of seeking clinician-investigator input for responses that required clinical interpretation. In addition, a manual of procedures for data collection and for the handling of special scenarios (e.g. readmissions or nosocomial infections) was circulated frequently to the sites throughout the study period. Second, all incoming data were manually reviewed by a data manager to identify missing or duplicate data, to verify that responses fell within accepted boundaries, and to assess for discrepant or conflicting responses. Third, data were reviewed in aggregate by the study team primarily to assess for inconsistencies and outliers by center. Data concerns prompted direct queries to the sites that were resolved prior to the final database freeze.

### Definitions

Digestive manifestations were divided into gastrointestinal symptoms and liver test abnormalities. Since anorexia is a common and non-specific symptom of viral illness, it was not considered a digestive manifestation in this study. Similarly, we did not include constipation because it has not previously been implicated as a symptom of acute or subacute viral infection. Therefore, the symptoms of interest in this study were diarrhea, nausea, vomiting, abdominal pain, gastrointestinal bleeding, dysphagia, and odynophagia. Patients were judged to have moderate-severe gastrointestinal symptoms when one or more of the following criteria were satisfied: 1) diarrhea with >4 bowel movements in any 24 hour period, 2) bloody diarrhea, 3) hematemesis, melena, or hematochezia, 4) abdominal computed tomography (CT) scan or endoscopic evaluation was performed, or 5) the gastroenterology consult service evaluated the patient. All other patients were considered to have mild symptoms.

Liver test abnormalities were defined as mild when the alanine aminotransferase level (ALT) or total bilirubin (TB) level was elevated between 1.5 and 3 times the upper limit of normal, moderate when there was >3 to 5 times elevation, and severe at >5 times elevation. Liver tests <1.5 times the upper limit were considered normal in this study. The upper limits of normal for ALT and TB were considered to be 45 units/L and 1.2 mg/dL respectively. Severe acute liver injury was defined as an ALT>1000 units/L with an international normalized ratio > 2 or a factor 5 level < 25%. For descriptive purposes, the proportion of patients with any liver test abnormality is reported, but only ALT and TB elevation were used in the analyses.

### Statistical analysis

Digestive manifestations were reported using descriptive statistics. Categorical variables were expressed as counts or percentages with 95% confidence intervals (95%CI); continuous variables were expressed as means with standard deviation (SD) or medians with interquartile range (IQR) depending on distribution. Liver test abnormality proportions were calculated using the full cohort as the denominator.

The association between digestive manifestations and the severity of COVID-19 was assessed using a multivariable logistic regression model which included the presence of gastrointestinal symptoms and liver test abnormalities as the independent variables of interest and which adjusted for pre-specified baseline covariates. The primary outcome was the composite endpoint of mechanical ventilation and/or death. Potential covariates that were considered for the model are listed in the **Supplementary Appendix**. If a univariable association with the outcome was observed (p<0.10), the covariate was considered for inclusion in the final regression model. We also explored potential interactions between included covariates and the primary independent variables. Only liver tests at admission were assessed, since hepatic injury during critical illness is consistently associated with multi-organ system failure and death regardless of etiology (22).

In exploratory analyses, the associations between digestive manifestations and intensive care unit admission, the need for vasopressor support, and hospital length of stay (modeled as a continuous variable) were assessed using a similar approach.

All analyses were conducted using SAS 9.4 (SAS Institute, Inc. Cary, North Carolina). The programming code for the final primary regression model is included in the **Supplementary Appendix**.

## Results

### Patients

From 15 April until 5 June 2020, data were collected from 1992 subjects across 36 centers. The median number of patients enrolled per participating institution was 51 (IQR 41-68). Characteristics of the study cohort are shown in **Table 1**. The average age was 60 years (SD 16.3); 57% were men; 42% were black/African American. Eighty-nine percent of patients had at least one non-digestive comorbidity; 9% had a pre-existing digestive disorder. Thirty-two percent of patients required mechanical ventilation and 19% died. The median hospital length of stay was 9 days (IQR 4-17). Thirty patients (1.5%) were still hospitalized at the end of the study period.

**Table 1:** Characteristics of the study cohort.

|  |  | All<br>N=1992 | 18-39<br>n=245 | 40-49<br>n=270 | 50-59<br>n=384 | 60-69<br>n=499 | 70-79<br>n=358 | 80-89<br>n=178 | >89<br>n=58 |
| --- | --- | --- | --- | --- | --- | --- | --- | --- | --- |
| **Demographics** |  |  |  |  |  |  |  |  |  |
| Age, mean (SD) |  | 60.1<br>(16.3) | 31.2<br>(6) | 45.2<br>(2.9) | 54.9<br>(2.7) | 64.4<br>(2.9) | 74.2<br>(2.9) | 83.8<br>(2.9) | >89*<br>(na) |
| Sex, n (%) | Male | 1128<br>(56.6) | 130<br>(53.1) | 175<br>(64.8) | 223<br>(58.1) | 294<br>(58.9) | 195<br>(54.5) | 86<br>(48.3) | 25<br>(43.1) |
|  | Female | 864<br>(43.4) | 115<br>(46.9) | 95<br>(35.2) | 161<br>(41.9) | 205<br>(41.1) | 163<br>(45.5) | 92<br>(51.7) | 33<br>(56.9) |
| Race**, n (%) | American Indian/Alaska Native | 5 (0.3) | 1 (0.4) | 0 (0) | 2 (0.5) | 1 (0.2) | 0 (0) | 1 (0.6) | 0 (0) |
|  | Asian | 70 (3.5) | 6 (2.4) | 11<br>(4.1) | 20<br>(5.2) | 18<br>(3.6) | 10<br>(2.8) | 3 (1.7) | 2<br>(3.4) |
|  | Black/African American | 842<br>(42.3) | 88<br>(35.9) | 119<br>(44.1) | 167<br>(43.5) | 223<br>(44.7) | 161<br>(45.1) | 68<br>(38.2) | 16<br>(27.6) |
|  | White | 732<br>(36.8) | 76<br>(31) | 72<br>(26.7) | 125<br>(32.6) | 186<br>(37.3) | 148<br>(41.5) | 89<br>(50) | 36<br>(62.1) |
|  | Multiple | 10 (0.5) | 1 (0.4) | 2 (0.7) | 3 (0.8) | 2 (0.4) | 1 (0.3) | 1 (0.6) | 0 (0) |
|  | Unknown | 332<br>(16.7) | 73<br>(29.8) | 66<br>(24.4) | 67<br>(17.4) | 69<br>(13.8) | 37<br>(10.4) | 16 (9) | 4<br>(6.9) |
| Ethnicity, n (%) | Hispanic or Latino | 290<br>(14.6) | 71<br>(29) | 67<br>(24.8) | 52<br>(13.5) | 51<br>(10.2) | 28<br>(7.8) | 16 (9) | 5<br>(8.6) |
|  | Not Hispanic or Latino | 1529<br>(76.8) | 146<br>(59.6) | 176<br>(65.2) | 300<br>(78.1) | 407<br>(81.6) | 301<br>(84.1) | 149<br>(83.7) | 50<br>(86.2) |
|  | Unknown | 173<br>(8.7) | 28<br>(11.4) | 27<br>(10) | 32<br>(8.3) | 41<br>(8.2) | 29<br>(8.1) | 13<br>(7.3) | 3<br>(5.2) |
| Health care worker, n (%) | Yes | 127<br>(6.4) | 26<br>(10.6) | 23<br>(8.5) | 41<br>(10.7) | 30 (6) | 3 (0.8) | 4 (2.2) | 0 (0) |
|  | No | 1653<br>(83) | 187<br>(76.3) | 212<br>(78.5) | 287<br>(74.7) | 410<br>(82.2) | 328<br>(91.6) | 172<br>(96.6) | 57<br>(98.3) |
|  | Unknown | 212<br>(10.6) | 32<br>(13.1) | 35<br>(13) | 56<br>(14.6) | 59<br>(11.8) | 27<br>(7.5) | 2 (1.1) | 1<br>(1.7) |
| **Prior history** |  |  |  |  |  |  |  |  |  |
| Cigarette smoking, n (%) | Current smoker | 126<br>(6.3) | 21<br>(8.6) | 15<br>(5.6) | 23 (6) | 40 (8) | 20<br>(5.6) | 7 (3.9) | 0 (0) |
|  | Ex-smoker | 569<br>(28.6) | 20<br>(8.2) | 37<br>(13.7) | 88<br>(22.9) | 165<br>(33.1) | 151<br>(42.2) | 85<br>(47.8) | 23<br>(39.7) |
|  | Non-smoker | 1175<br>(59) | 195<br>(79.6) | 201<br>(74.4) | 253<br>(65.9) | 269<br>(53.9) | 161<br>(45) | 71<br>(39.9) | 25<br>(43.1) |
|  | Unknown | 122<br>(6.1) | 9 (3.7) | 17<br>(6.3) | 20<br>(5.2) | 25 (5) | 26<br>(7.3) | 15<br>(8.4) | 10<br>(17.2) |
| Alcoholism, n (%) | Current | 169<br>(8.5) | 26<br>(10.6) | 29<br>(10.7) | 37<br>(9.6) | 41<br>(8.2) | 26<br>(7.3) | 6 (3.4) | 4<br>(6.9) |
|  | Prior | 102<br>(5.1) | 7 (2.9) | 10<br>(3.7) | 23 (6) | 24<br>(4.8) | 24<br>(6.7) | 11<br>(6.2) | 3<br>(5.2) |
|  | No | 1500<br>(75.3) | 194<br>(79.2) | 204<br>(75.6) | 283<br>(73.7) | 387<br>(77.6) | 260<br>(72.6) | 131<br>(73.6) | 41<br>(70.7) |
|  | Unknown | 221<br>(11.1) | 18<br>(7.3) | 27<br>(10) | 41<br>(10.7) | 47<br>(9.4) | 48<br>(13.4) | 30<br>(16.9) | 10<br>(17.2) |
| Obesity, n (%) |  | 968<br>(48.6) | 151<br>(61.6) | 151<br>(55.9) | 205<br>(53.4) | 264<br>(52.9) | 147<br>(41.1) | 46<br>(25.8) | 4<br>(6.9) |
| Body mass index***, mean (SD) |  | 31.5<br>(8.5) | 35.2<br>(10.3) | 33.6<br>(8.9) | 32.5<br>(9.3) | 31.8<br>(7.5) | 29.1<br>(6.2) | 27.1<br>(6.4) | 23.9<br>(4.1) |
| Charlson Comorbidity Index, median (IQR) |  | 1 (0, 2) | 0 (0, 1) | 0 (0, 1) | 1 (0, 2) | 1 (0, 3) | 2 (1, 3) | 2 (1, 3) | 2 (1, 3) |
| Diabetes, n (%) |  | 722<br>(36.2) | 50<br>(20.4) | 75<br>(27.8) | 133<br>(34.6) | 221<br>(44.3) | 152<br>(42.5) | 79<br>(44.4) | 12<br>(20.7) |
| Hypertension, n (%) |  | 1245<br>(62.5) | 55<br>(22.4) | 121<br>(44.8) | 223<br>(58.1) | 362<br>(72.5) | 280<br>(78.2) | 156<br>(87.6) | 48<br>(82.8) |
| Cardiac disease, n (%) |  | 435<br>(21.8) | 12<br>(4.9) | 14<br>(5.2) | 55<br>(14.3) | 135<br>(27.1) | 117<br>(32.7) | 74<br>(41.6) | 28<br>(48.3) |
| Pulmonary disease, n (%) |  | 414<br>(20.8) | 46<br>(18.8) | 52<br>(19.3) | 74<br>(19.3) | 116<br>(23.2) | 75<br>(20.9) | 40<br>(22.5) | 11<br>(19) |
| Active/current malignancy,<br>excluding non-melanoma skin<br>cancer, n (%) |  | 123<br>(6.2) | 6 (2.4) | 7 (2.6) | 24<br>(6.3) | 42<br>(8.4) | 34<br>(9.5) | 7 (3.9) | 3<br>(5.2) |
| Immunocompromised, n (%) |  | 265<br>(13.3) | 39<br>(15.9) | 30<br>(11.1) | 59<br>(15.4) | 76<br>(15.2) | 51<br>(14.2) | 7 (3.9) | 3<br>(5.2) |
| Luminal gastrointestinal disease<br>(non-malignant), n (%) |  | 84 (4.2) | 5 (2) | 7 (2.6) | 16<br>(4.2) | 25 (5) | 22<br>(6.1) | 8 (4.5) | 1<br>(1.7) |
| Pancreaticobiliary disease, n (%) |  | 56 (2.8) | 2 (0.8) | 8 (3) | 11<br>(2.9) | 15 (3) | 17<br>(4.7) | 3 (1.7) | 0 (0) |
| Chronic liver disease, n (%) |  | 55 (2.8) | 2 (0.8) | 8 (3) | 14<br>(3.6) | 17<br>(3.4) | 10<br>(2.8) | 4 (2.2) | 0 (0) |
| **COVID-19 symptoms** |  |  |  |  |  |  |  |  |  |
| Fever (subjective or objective), n<br>(%) |  | 1537<br>(77.2) | 209<br>(85.3) | 222<br>(82.2) | 303<br>(78.9) | 390<br>(78.2) | 254<br>(70.9) | 122<br>(68.5) | 37<br>(63.8) |
| Cough, n (%) |  | 1476<br>(74.1) | 197<br>(80.4) | 220<br>(81.5) | 301<br>(78.4) | 362<br>(72.5) | 248<br>(69.3) | 112<br>(62.9) | 36<br>(62.1) |
| Shortness of breath, n (%) |  | 1403<br>(70.4) | 186<br>(75.9) | 204<br>(75.6) | 279<br>(72.7) | 359<br>(71.9) | 232<br>(64.8) | 113<br>(63.5) | 30<br>(51.7) |
| Fatigue or subjective weakness, n<br>(%) |  | 851<br>(42.7) | 86<br>(35.1) | 105<br>(38.9) | 173<br>(45.1) | 216<br>(43.3) | 167<br>(46.6) | 77<br>(43.3) | 27<br>(46.6) |
| Myalgia, n (%) |  | 580<br>(29.1) | 97<br>(39.6) | 111<br>(41.1) | 127<br>(33.1) | 141<br>(28.3) | 73<br>(20.4) | 24<br>(13.5) | 7<br>(12.1) |
| **COVID-19 treatments** |  |  |  |  |  |  |  |  |  |
| Hydroxychloroquine/Chloroquine, n (%) |  | 1036<br>(52) | 115<br>(46.9) | 137<br>(50.7) | 207<br>(53.9) | 280<br>(56.1) | 185<br>(51.7) | 95<br>(53.4) | 17<br>(29.3) |
| Remdesivir, n (%) |  | 109<br>(5.5) | 13<br>(5.3) | 20<br>(7.4) | 21<br>(5.5) | 37<br>(7.4) | 9 (2.5) | 7 (3.9) | 2<br>(3.4) |
| Convalescent plasma, n (%) |  | 37 (1.9) | 4 (1.6) | 4 (1.5) | 9 (2.3) | 11<br>(2.2) | 7 (2) | 2 (1.1) | 0 (0) |
| Glucocorticoids, n (%) |  | 240<br>(12) | 23<br>(9.4) | 31<br>(11.5) | 39<br>(10.2) | 69<br>(13.8) | 54<br>(15.1) | 22<br>(12.4) | 2<br>(3.4) |
| Tocilizumab, n (%) |  | 109<br>(5.5) | 19<br>(7.8) | 16<br>(5.9) | 25<br>(6.5) | 32<br>(6.4) | 9 (2.5) | 7 (3.9) | 1<br>(1.7) |
| **COVID-19 outcomes** |  |  |  |  |  |  |  |  |  |
| Hospital length of stay, days, median (IQR) |  | 9 (4,17) | 6<br>(3,11) | 7<br>(4,15) | 8<br>(4,17) | 11<br>(5,23) | 10<br>(6,19) | 10.5<br>(6,18) | 8<br>(5,14) |
| Intensive care unit admission, n (%) |  | 878<br>(44.1) | 80<br>(32.7) | 101<br>(37.4) | 157<br>(40.9) | 269<br>(53.9) | 178<br>(49.7) | 74<br>(41.6) | 19<br>(32.8) |
| Mechanical ventilation, n (%) |  | 646<br>(32.4) | 53<br>(21.6) | 77<br>(28.5) | 113<br>(29.4) | 211<br>(42.3) | 134<br>(37.4) | 52<br>(29.2) | 6<br>(10.3) |
| Vasopressor support, n (%) |  | 546<br>(27.4) | 40<br>(16.3) | 58<br>(21.5) | 94<br>(24.5) | 179<br>(35.9) | 125<br>(34.9) | 44<br>(24.7) | 6<br>(10.3) |
| Death, n (%) |  | 375<br>(18.8) | 16<br>(6.5) | 20<br>(7.4) | 44<br>(11.5) | 93<br>(18.6) | 102<br>(28.5) | 66<br>(37.1) | 34<br>(58.6) |

\*Age was not collected for patients >89 years old
\*\*1 missing
\*\*\*172 missing

### Prevalence, spectrum, and severity of gastrointestinal symptoms

Overall, 1052 patients (53%, 95%CI 51-55%) experienced at least one gastrointestinal symptom at any time during their illness (**Figure 1**). Of these, 227 patients (11%, 95%CI 10-13%) experienced 3 or more gastrointestinal symptoms. The most common symptoms were diarrhea (34%, 95%CI 32-36%), nausea (27%, 95%CI 25-29%), vomiting (16%, 95%CI 14-17%), and abdominal pain (11%, 95%CI 10-13%). The prevalence of gastrointestinal symptoms and their distribution did not differ substantively after excluding patients with preexisting gastrointestinal luminal and pancreaticobiliary diseases. The overall proportion decreased to 47% (95%CI 44-50%) after excluding patients who were known to have received COVID-19 treatments that may be associated with gastrointestinal side-effects, such as hydroxychloroquine or remdesivir (**Supplementary Appendix**). The prevalence of gastrointestinal symptoms varied across sites (**Supplementary Appendix**).

**Figure 1:**
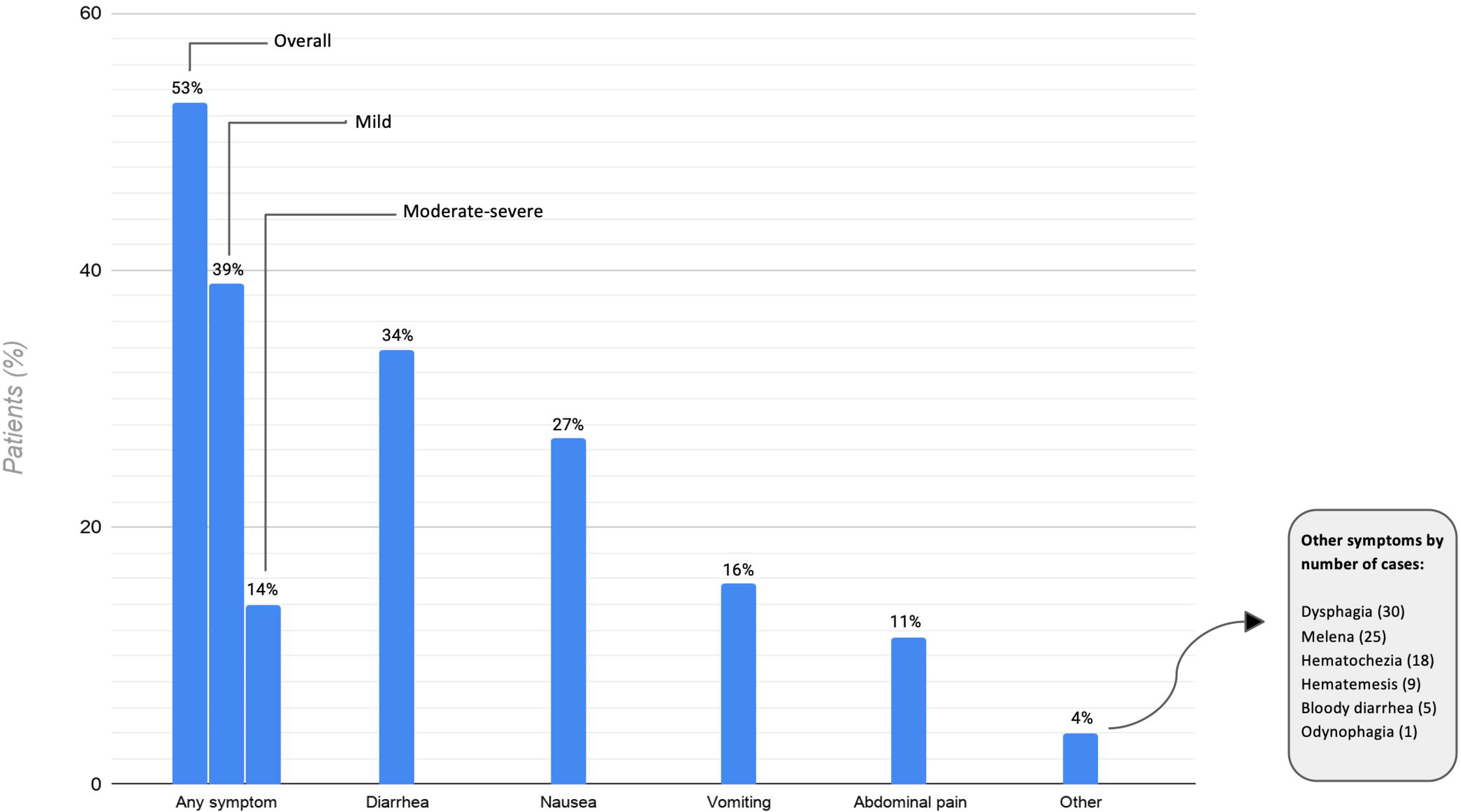
Gastrointestinal symptoms in patients hospitalized with COVID-19.

Gastrointestinal symptoms preceded other COVID-19 symptoms in 13% of cases, started concurrently in 44%, and followed the other COVID-19 symptoms in 42%. In 7 patients (0.4%), gastrointestinal symptoms were the only manifestation of COVID-19.

In total, 74% of patients (781 of 1052) with gastrointestinal symptoms were judged to have mild symptoms according to our criteria. Among the 271 patients with moderate-severe symptoms, 21% had diarrhea with >4 bowel movements per 24 hours, 2% had bloody diarrhea, 18% had gastrointestinal hemorrhage, 63% underwent abdominal CT scan (159 patients) and/or endoscopic examination (19 patients), and 23% were evaluated by the gastroenterology consult service. Gastrointestinal symptoms were judged to be less prominent than other COVID-19 symptoms in 73% of patients, equally prominent in 20% of patients, and more prominent in 6%.

### Prevalence, spectrum, and severity of liver test abnormalities

Liver tests were available in 1712 patients (86%) at presentation. At the time of admission, 554 patients (28% of the full cohort, 95%CI 26-30%) had at least one abnormal liver test (**Figure 2**). Of these, 215 (11%, 95%CI 11-14%) had an abnormal ALT or TB. Among patients with an abnormal ALT or TB, 77% (95%CI 71-82%) had mild elevation, 17% (95%CI 12-22%) had moderate elevation, and 6% (95%CI 3-9%) had severe elevation. Median abnormal liver test values are presented in **Figure 2**.

**Figure 2:**
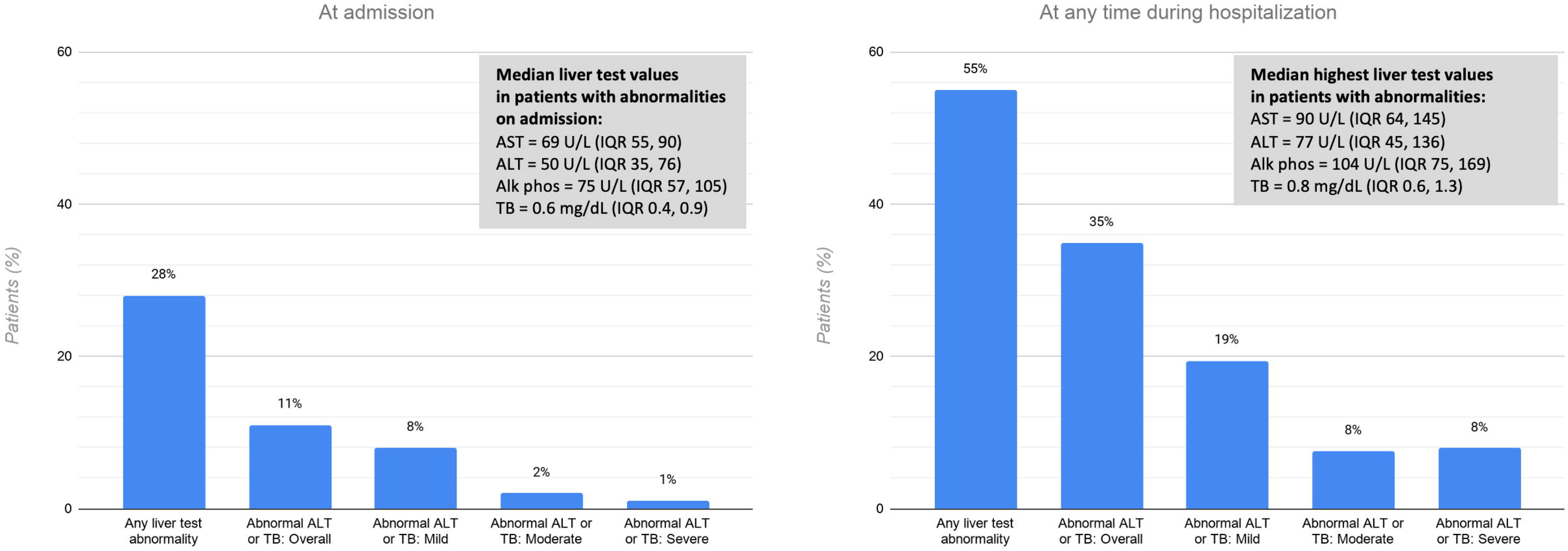
Liver test abnormalities in patients hospitalized with COVID-19, at admission and at any time during hospitalization. Proportions were calculated using the full cohort as the denominator (N=1992). AST – aspartate aminotransferase: ALT – alanine aminotransferase; Alk phos – alkaline phosphatase; TB – total bilirubin; IQR – interquartile range

Liver tests were available in 1890 patients (95%) at any time during hospitalization. Among those with normal liver tests at admission, 548 patients developed at least one abnormality during their hospitalization; of these, 480 (24%, 95%CI 22-26) developed an abnormal ALT or TB. In total, 695 (35%, 95%CI 33-37%) had an elevated ALT or TB; 56% (95%CI 52-59) of these had mild elevation, 22% (95%CI 18-25) had moderate elevation, and 23% (95%CI 19-26) had severe elevation. Twenty-three patients (1%) developed an ALT level in excess of 1000 U/L during the hospitalization and 5 of these (0.3%) met criteria for severe acute liver injury. No patients had an ALT >1000 U/L on admission. The overall proportion, pattern, and severity of abnormal ALT or TB levels did not differ after excluding patients with prior liver disease or those known to have received COVID-19 treatments that can cause hepatotoxicity (**Supplementary Appendix**).

### Association between digestive manifestations and severe COVID-19 outcomes

After adjusting for potential confounders, the presence of gastrointestinal symptoms was not associated with the primary composite endpoint of mechanical ventilation and/or death (odds ratio [OR] 0.93, 95%CI 0.76-1.15). Similarly, the presence of mild (OR 1.05, 95%CI 0.73-1.49), moderate (OR 1.14, 95%CI 0.56-2.32), or severe (OR 1.89, 95%CI 0.57-6.30) liver test abnormalities on admission was not associated with the primary endpoint. In the exploratory analyses, there was no association between digestive manifestations and intensive care unit admission, the need for vasopressor support, or hospital length of stay.

## Discussion

In this large and geographically diverse cohort of patients hospitalized with COVID-19 in North America, 53% of patients experienced at least one gastrointestinal symptom and 35% developed an abnormal ALT or TB level at some point during their illness. The majority of gastrointestinal symptoms and hepatic abnormalities were mild in nature. The presence of gastrointestinal symptoms at any time or liver test abnormalities on admission were not associated with mechanical ventilation or death.

While liver test abnormalities are objective, the assessment of gastrointestinal manifestations in COVID-19 is limited by uncertainties in symptom attribution and ascertainment. In this study, we considered any symptom without a clear alternative explanation (e.g. abdominal pain due to a known post-operative complication) as potentially attributable to COVID-19. This approach likely overestimated prevalence since some symptoms may have been related to other factors such as medication effect (e.g. diarrhea or nausea) or critical illness (e.g. gastrointestinal hemorrhage or dysphagia). Indeed, a subgroup analysis that excluded patients who were documented to have received COVID-19 treatments that could cause digestive side-effects showed a small decrease in the prevalence of gastrointestinal symptoms (47%, 95%CI 44-50% vs. 53%, 95%CI 51-55% overall). Similarly, manual review of endoscopy cases revealed that approximately half were performed to address events related to critical illness (e.g. feeding tube placement) rather than direct viral injury (data not shown). Conversely, since the focus of care in hospitalized patients with COVID-19 is typically the pulmonary process, it is possible that gastrointestinal symptoms were under-reported and/or under-documented. Along these lines, abdominal imaging and endoscopy – criteria we used to determine the severity of gastrointestinal symptoms in this study – have been employed judiciously during the pandemic to minimize in-hospital exposure, perhaps underestimating the significance of symptoms. Nevertheless, our findings provide valuable information on the overall burden of digestive manifestations and associated resource utilization in patients hospitalized with COVID-19, whether due to direct viral effect, treatment of the infection, or a consequence of related systemic illness.

The prevalence of digestive manifestations in this study was consistent across the majority of participating institutions and in line with that observed in other Western studies (16,17). This is in contrast to much lower proportions of gastrointestinal symptoms reported in studies from China (9,10). This difference may be because the Chinese experience largely reflects the early phase of the pandemic, prior to widespread recognition of digestive symptoms as a frequent consequence of COVID-19. These early studies aimed to better understand the overall illness, whereas more recent studies from the West have focused specifically on identifying gastrointestinal symptoms. Alternatively, variable disease expression between patient populations as a result of genetic or epigenetic factors or prevalent virus mutations (23) may explain the difference in proportions and deserve more attention.

The effect of digestive involvement on pulmonary and systemic illness through gut-lung crosstalk or other unknown mechanisms is of major potential importance. For example, microbiome-driven interferon signatures have been shown to suppress viral replication in the lung and can be disrupted by gut dysbiosis in experimental models of influenza infection (7). Moreover, the small intestine comprises a rich immune apparatus, the dysregulation of which by SARS-CoV-2 could potentiate or even drive systemic inflammatory response (8). Our findings, however, do not support such a hypothesis given the lack of association between gastrointestinal symptoms and overall severity of illness. Additional research is necessary to elucidate whether the gut-lung axis is influential in this disease independent of gastrointestinal symptoms.

Our findings are consistent with prior reports demonstrating that liver tests abnormalities are common in COVID-19. Fifty-five percent of patients in this cohort had an elevated liver test at some point during their illness, including many with abnormal aminotransferases, raising the possibility of hepatocyte injury due to SARS-CoV-2. However, the large majority had ALT or TB levels less than 5 times the upper limit of normal and only 23 patients had an ALT >1000 units/L, which was never present at the time of presentation, suggesting that clinically important liver injury in COVID-19 is uncommon. Future mechanistic investigation will be necessary to better understand whether infection leads to direct hepatic injury.

The findings of this study should be interpreted in the context of several limitations, some of which are inherent to observational research on COVID-19. As highlighted above, symptom attribution and ascertainment were influenced by several factors related to conducting research during a pandemic, such as retrospective data collection and reliance on medical records review rather than direct patient interviews. Further, validated definitions for gastrointestinal symptom severity in COVID-19 are not available and thus we devised criteria that we believe reasonably reflect disease severity in terms of patient suffering and resource utilization. Alternative definitions of severity may have led to varying interpretations of the findings. Some of these limitations are mitigated by the large and geographically diverse sample, highly systematic approach to patient selection, and multi-layered and rigorous strategy to ensure the veracity of collected data. It is also important to consider that this study was restricted to hospitalized patients and thus does not reflect the prevalence and significance of digestive manifestations in outpatients with COVID-19.

In summary, among patients hospitalized with COVID-19, gastrointestinal symptoms and liver test abnormalities were common, but the majority were mild in nature and their presence was not associated with worse clinical outcomes.

## Data Availability

All authors had access the data

## References

1) Xiao F, Tang M, Zheng X, Liu Y, et al. Evidence for Gastrointestinal Infection of SARS-CoV-2. Gastroenterol 2020;158:1831-1833.

2) Du M, Cai G, Chen F, et al. Multi-omics Evaluation of Gastrointestinal and Other Clinical Characteristics of SARS-CoV-2 and COVID-19. Gastroenterol 2020;158:2298-230.

3) ACE2 angiotensin I converting enzyme 2 [Homo sapiens (human)]. Gene ID: 59272. (https://www.ncbi.nlm.nih.gov/gene/59272).

4) Han C, Duan C, Zhang S, et al. Digestive Symptoms in COVID-19 Patients with Mild Disease Severity: Clinical Presentation, Stool Viral RNA Testing, and Outcomes. Am J Gastroenterol 2020;115:916-923.

5) Wu Y, Guo C, Tang L, et al. Prolonged presence of SARS-CoV-2 viral RNA in faecal samples. Lancet Gastroenterol Hepatol 2020;5:434-435.

6) Bradley KC, Finsterbusch K, Schnepf D, et al. Microbiota-driven tonic interferon signals in lung stromal cells protect from influenza virus infection. Cell Rep 2019;28:245-256.

7) Marsland BJ, Trompette A, Gollwitzer ES. The Gut-Lung Axis in Respiratory Disease. Ann Am Thorac Soc 2015;12 Suppl 2:S150-6.

8) Mönkemüller K, Fry L, Rickes S. COVID-19, Coronavirus, SARS-CoV-2 and the small bowel. Rev Esp Enferm Dig 2020;112:383-388.

9) Guan WJ, Ni ZY, Hu Y, et al. Clinical Characteristics of Coronavirus Disease 2019 in China. N Engl J Med 2020;382:1708-1720.

10) Huang C, Wang Y, Li X, et al. Clinical features of patients infected with 2019 novel coronavirus in Wuhan, China. Lancet 2020;395:497-506.

11) Jin X, Lian JS, Hu JH, et al. Epidemiological, clinical and virological characteristics of 74 cases of coronavirus-infected disease 2019 (COVID-19) with gastrointestinal symptoms. Gut 2020;69:1002-1009.

12) Sultan S, Altayar O, Siddique S, et al. AGA Institute Rapid Review of the GI and Liver Manifestations of COVID-19, Meta-Analysis of International Data, and Recommendations for the Consultative Management of Patients with COVID-19. Gastroenterology (2020), doi: https://doi.org/10.10537j.gastro.2020.05.001.

13) Parasa S, Desai M, Thoguluva Chandrasekar V, et al. Prevalence of Gastrointestinal Symptoms and Fecal Viral Shedding in Patients With Coronavirus Disease 2019: A Systematic Review and Meta-analysis. JAMA Network Open 2020;3(6):e2011335. doi: 10.1001/jamanetworkopen.2020.11335.

14) D’Amico F, Baumgart DC, Danese S, Peyrin-Biroulet L. Diarrhea during COVID-19 infection: pathogenesis, epidemiology, prevention and management. Clin Gastroenterol and Hepatol 2020 April 8. doi: https://doi.org/10.1016/j.cgh.2020.04.001.

15) Aroniadis OC, DiMaio CJ, Dixon RE, et al. Current Knowledge and Research Priorities in the Digestive Manifestations of COVID-19. Clin Gastroenterol Hepatol. 2020 Apr 22. pii: S1542-3565(20)30536-X. doi: 10.1016/j.cgh.2020.04.039. [Epub ahead of print]

16) Redd WD, Zhou JC, Hathorn KE, et al. Prevalence and Characteristics of Gastrointestinal Symptoms in Patients with SARS-CoV-2 Infection in the United States: A Multicenter Cohort Study. Gastroenterology. 2020 Apr 22. pii: S0016-5085(20)30564-3. doi: 10.1053/j.gastro.2020.04.045. [Epub ahead of print]

17) Hajifathalian K, Krisko T, Mehta A, et al. Gastrointestinal and Hepatic Manifestations of 2019 Novel Coronavirus Disease in a Large Cohort of Infected Patients From New York: Clinical Implications. Gastroenterology. 2020 May 7:S0016-5085(20)30602-8. doi: 10.1053/j.gastro.2020.05.010. Online ahead of print.

18) Zhang C, Shi L, Wang FS. Liver injury in COVID-19: management and challenges. Lancet Gastroenterol Hepatol 2020;5:428-430.

19) Pan L, Yang P, Sun Y, et al. Clinical characteristics of COVID-19 patients with digestive symptoms in Hubei, China: a descriptive, cross-sectional, multicenter study. Am J Gastroenterol 2020;115(5):766-773.

20) Wei X-S, Wang X, Niu Y-R, et al. Diarrhea is associated with prolonged symptoms and viral carriage in COVID-19. Clinical Gastroenterology and Hepatology. 2020 doi: https://doi.org/10.10167j.cgh.2020.04.030.

21) Mao R, Qiu Y, He JS. Manifestations and Prognosis of Gastrointestinal and Liver Involvement in Patients With COVID-19: A Systematic Review and Meta-Analysis Lancet Gastroenterol Hepatol 2020 May 12. doi: 10.1016/S2468-1253(20)30126-6.

22) Lone NI, Walsh TS. Impact of Intensive Care Unit Organ Failures on Mortality During the Five Years After a Critical Illness. Am J Respir Crit Care Med 2012;186:640-7.

23) Coppée F, Lechien JR, Declèves AE, et al. Severe acute respiratory syndrome coronavirus 2: virus mutations in specific European populations. New Microbes New Infect 2020;36:100696.

